# SPATIAL HETEROGENEITY OF NEIGHBORHOOD-LEVEL WATER AND SANITATION ACCESS IN INFORMAL URBAN SETTLEMENTS: A CROSS-SECTIONAL CASE STUDY IN BEIRA, MOZAMBIQUE

**DOI:** 10.1101/2022.01.25.22269649

**Authors:** Courtney Victor, Denisse Vega Ocasio, Zaida A. Cumbe, Joshua V. Garn, Sydney Hubbard, Magalhaes Mangamela, Sandy McGunegill, Rassul Nalá, Jedidiah S. Snyder, Karen Levy, Matthew C Freeman

**Affiliations:** Gangarosa Department of Environmental Health, Emory University, 1518 Clifton Rd. NE, Atlanta, GA 30322, USA; WEConsult, Maputo, Mozambique; School of Community Health Sciences, University of Nevada, 1664 N. Virginia Street, Reno, NV, 89557, USA; AURA – Autoridade Reguladora de Água, Former Executive Secretary of the Water Regulation Council (CRA), currently (AURA, IP), Water Regulatory Authority, Public Institute; Department of Environmental and Occupational Health Sciences, School of Public Health, University of Washington, 3980 15^th^ Avenue NE, Seattle, Washington, USA; INS – Instituto Nacional de Saúde, Ministério de Saúde, EN1 Bairro da Vila-parcela № 3943, Distrito de Marracuene, Maputo, República de Moçambique

**Keywords:** water, sanitation, hygiene, spatial heterogeneity, Mozambique

## Abstract

Rapid urbanization, resulting in population growth within informal settlements, has worsened exclusion and inequality in access to water and sanitation (WASH) services in the poorest and most marginalized communities. In this study, we describe the heterogeneity in water service satisfaction and WASH access in low-income, peri-urban neighborhoods of Beira, Mozambique, and examine whether this heterogeneity can be explained by distance to water distribution mains. Using spatial statistics and regression analyses, we identified statistical spatial heterogeneity in household WASH access, as well as consumer-reported satisfaction with water services (services, pressure, quality, and sufficient quantity). We found that as distance from the water main increased, both access to an improved water source at the household and satisfaction with water pressure decreased, controlling for household density and socioeconomic status. The odds of a household having access to a water source at the household or on the compound decreased with every 100-meter increase in distance from a water main pipe (odds ratio [OR] 0.87, 95% confidence interval [CI]: 0.82, 0.92). Satisfaction with water services also decreased with every 100-meter increase in distance from a water main pipe (OR: 0.80; 95% CI: 0.69, 0.94). Findings from this study highlight the unequal household access to water and sanitation in urban informal settlements, even within low-income neighborhoods. Describing this heterogeneity of access to water services, sanitation, and satisfaction – and the factors influencing them - can inform stakeholders and guide the development of infrastructural solutions to reduce water access inequities within urban settings.

## Introduction

Rapid urbanization has brought new challenges and urgency concerning access to improved water and sanitation services.(1) In sub-Saharan Africa, nearly 60% of people living in cities reside in informal settlements, defined as urban areas where residents lack access to basic public services, goods and amenities, and formal and secure tenure.(2) In many low- and middle-income countries (LMICs), population growth within informal settlements has overstretched existing water supply and sewerage networks and presented difficulty for populations to access distribution networks.(3) Despite improvements in water and wastewater systems, unequal access to basic services remains a global risk factor for exposure to fecal pathogens.(4, 5) In 2020, 771 million people lacked basic drinking water service, and 1.7 billion people lacked basic sanitation service.(6) In 2017, diarrheal diseases accounted for almost 1.6 million deaths worldwide, and they were the second leading cause of mortality in children under the age of five.(7, 8) Limited access to improved sanitation systems contributes to outbreaks of waterborne and respiratory diseases, including COVID-19, as well as malnutrition and impaired educational outcomes and social and economic development.(6, 9-12)

Urbanization has particularly exacerbated exclusion and inequality in access to WASH services for the poorest and most marginalized families. As of 2018, 4.2 billion people were living in urban areas globally; of those, about 300 million were children.(13, 14) In sub-Saharan Africa, the fastest urbanizing region globally, the mismatch in investment, expansion, and maintenance in urban water and sanitation services has resulted in delays in the expansion of services to informal settlement areas, as well as reduced operational sustainability of these services.(15, 16) Consequently, many residents from unplanned settlements lack access to basic services such as water supply and sanitation.(3)

Access to water and sanitation services within urban informal settlements is typically poor, but not uniformly so. While limited access to water and sanitation services is more pronounced in informal and peri-urban settlements,(17, 18) there is limited data on heterogeneity *within* urban informal settlements, and a lack of analysis on what drives these underlying inequities. Among the factors known to contribute to this problem are challenges with distribution network engineering in informal, unplanned neighborhoods, issues of housing and network expansion planning, and failure of public policy to provide satisfactory solutions to both address and ensure access to safe and continuous water supply.(17-19) While significant progress has been made towards addressing inequities in access to improved water and sanitation facilities, global measures of coverage overestimate those with reliable, high-quality services.(5, 17, 20) Equitable access to safe water and sanitation systems is achieved as long as the principles of operational sustainability are upheld (21). These include both the functionality of the systems themselves and the household’s experience of quality services over time. (21) Therefore, understanding local variability in water and sanitation access reflected by level of satisfaction with services, and the factors influencing heterogeneous access in informal settlements, is important for designing improvements to expand reliable, high quality coverage and access in these settings.

Relative to other African countries of similar income, Mozambique has experienced fewer and unequal improvements in access to basic water and sanitation services since 2010. Access to at least basic water sources in Mozambique was approximately 63% in 2020.(5) In the same year, access to at least basic sanitation services was around 37% in Mozambique, less than half the access in the Democratic Republic of the Congo, and lower than Nigeria, and Rwanda.(5) Water and sanitation coverage rates in Mozambique are higher in urban areas than rural areas, yet access to at least basic facilities is unevenly distributed across the country.(5, 22). It is one of 15 countries with a gap in subnational basic sanitation coverage of greater than 50%, and the ratio for basic drinking water coverage comparing the richest wealth quintiles to the poorest wealth quintiles was 1.8 in 2019.(5) The slow and unequal increase in facilities in urban areas of Mozambique has been attributed to limited resources and funding to maintain or improve current infrastructure, limited emphasis on service and water quality, and challenges related to informal settlements infrastructures.(5, 22, 23)

In this study, we describe heterogeneity in water access in low-income, peri-urban neighborhoods of Beira, Mozambique. We combine spatial statistical methods and regression analyses to investigate differences and inequities in access to improved drinking water, improved sanitation, and consumer-reported satisfaction with water services. We explore whether factors such as distance to water main pipes, socioeconomic status (SES), and household density influence access to improved sanitation and water services, and how water satisfaction varies across the city of Beira. The results of these analyses provide finer detail about access to improved water and sanitation services that facilitate our understanding of the drivers of variability of access to improved water and sanitation facilities, even within urban informal settlements. Data on intra-neighborhood heterogeneity could support local service providers and stakeholders in planning efforts to expand and improve service delivery to those who most need it.

## Methods

The purpose of this study was to characterize water and sanitation access and satisfaction in low-income, peri-urban neighborhoods of Beira City. We asked the following research questions: 1) What is the spatial heterogeneity of access to improved water and sanitation and satisfaction with water services within low-income neighborhoods? And 2) Does distance to water distribution mains drive access to a household connection to a piped water source or satisfaction with water services? Data were derived from a population-based survey conducted in 14 low-income areas from central Beira City. This survey was part of formative research for a parent study, titled “*Pesquisa sobre o Acesso à Água e a Saúde Infantil em Moçambique (PAASIM)*”, designed to assess the health impacts of piped water supply on young children in Beira.

### Study site

Beira, a coastal city in Sofala Province with a high-water table, at the mouth of the Púnguè River, is the second-largest city in Mozambique, with a population of around 530,000 individuals (Figure 1). Mozambique’s water and sanitation sector is overseen by the National Directorate of Water Supply and Sanitation (DNAAS), Water-Supply Asset Holding and Investment Fund (FIPAG) and the Water and Sanitation Infrastructure Administration (AIAS). FIPAG, an entity of the Delegated Management Framework for Water Supply (QGD) in Mozambique, is responsible for the management of assets, and both the public and private investment programs in the urban water supply systems. FIPAG is also responsible for promoting autonomous, efficient, and profitable management of the water system, namely through the transfer of operations to private operators. (23, 24) Economic regulation and consumer protection in the service provision is carried out by the Water Regulatory Authority (CRA), which as of February of 2019, became Water Regulatory Authority (AURA. IP). FIPAG faces various challenges in providing water service in Beira and increasing coverage in informal urban settlements. The mandate of AIAS is to promote the autonomous, efficient, and financially sustainable management of public water supply and sanitation systems. They are responsible for managing the assets of secondary public water distribution and sanitation systems.

**Figure 1:**
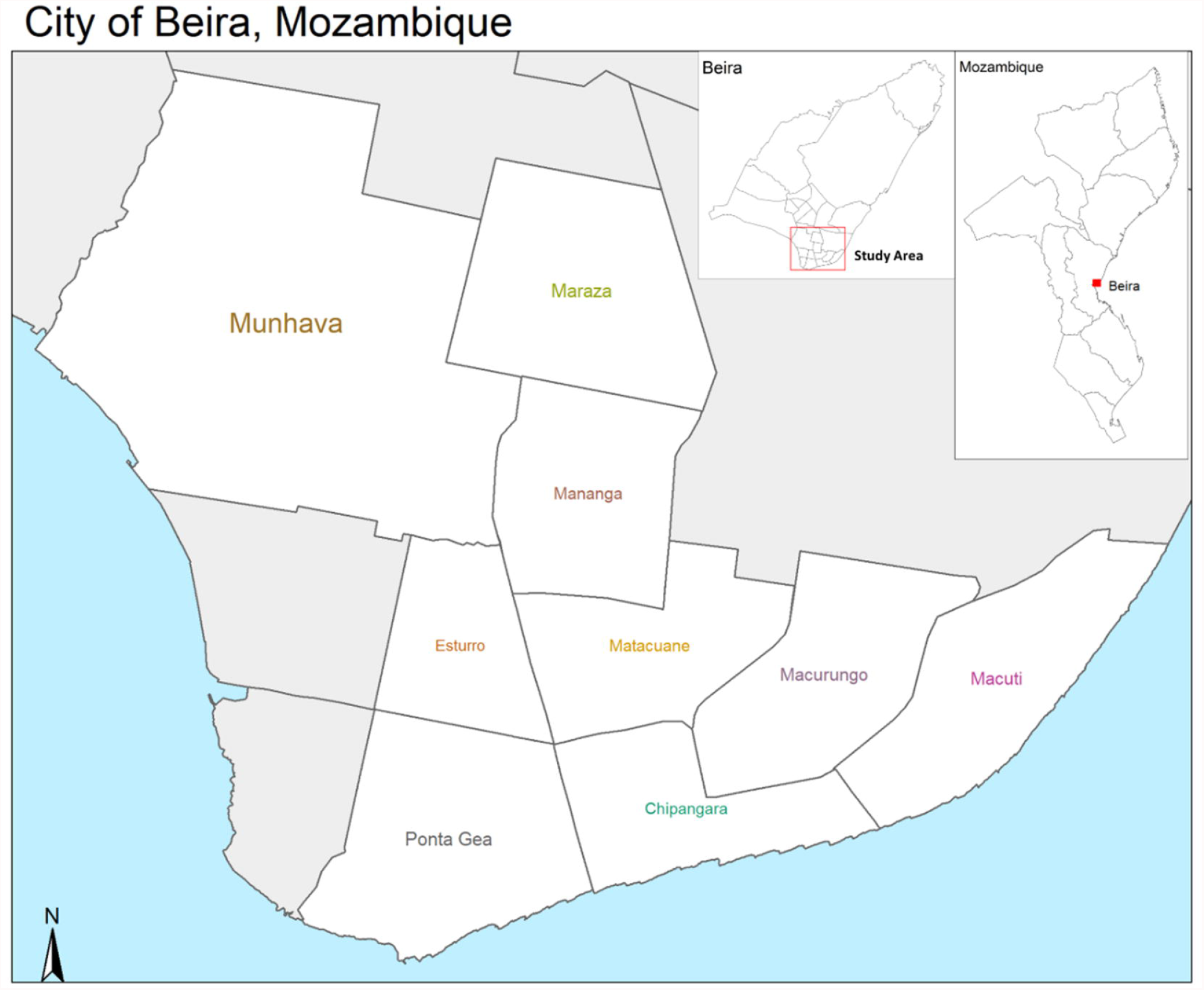
Map of the area of the city of Beira, Mozambique where the study was conducted. A subset of low-income, high-density neighborhoods were chosen for the population-based survey analyzed in this study.

### Neighborhood Selection and Household sampling scheme

We selected a set of sub-neighborhoods within the city center, primarily based on their characteristics as containing low-income (see definition in *Predictor Variables*), high-density, peri-urban housing, and specifically including areas that had received or were targeted to receive a new water network (intervention areas) and similar comparison areas, which we are examining in the PAASIM study (Figure 1). Sub-neighborhood boundaries were delineated along natural boundaries such as roads or waterways and based on maps received from FIPAG showing areas that were scheduled to receive or not receive the intervention. We aimed to target areas inhabited by predominantly low-income residents. We then used a probability-proportionate-to-size sampling scheme to select a representative sample of single-story households from these areas.

The number of households and population density of each sub-neighborhood was approximated through household density estimates using Google Earth satellite imagery. We applied a random grid method, where a grid was placed over an area, and a random selection of squares were selected from that area. Two researchers manually counted households in the randomly selected squares, and the number of houses per unit was extrapolated across unsampled squares. Household density estimates were used to determine proportional sampling (using household counts instead of population), where the probability of a household being selected into the study was proportional to the household density of the neighborhood.

Enumerators used an interactive map of study neighborhoods to select a grid of their assigned segment (sub-neighborhood) to begin sampling. At this point, the enumerator randomly selected the first house using a random number generator between 1 and 19. Enumerators then systematically sampled every 19^th^ household until all households had been counted in the sub-neighborhood, to provide approximately a 5% proportional sample. The enumerator recorded survey results for sampled households that were abandoned (n=20), had no eligible adult respondent available at the time of the survey (n=143), or the respondent refused to consent to participate in the survey (n=47) and moved on to the next household. The geolocation of sampled households was uploaded to the interactive map daily to ensure that areas were not missed or skipped. Community members were assigned by the sub-neighborhood head to help guide the enumerators to areas if there was no clear path. Vertical slums and two-story households were excluded from the study due to logistical challenges with sampling and conducting surveys. Further, observations during our study site selection suggested that these households were not representative of low-income peri-urban neighborhoods.

### Data Collection

Data were collected from November to December 2019. The survey instrument consisted of several modules, including questions regarding household demographics, assets and wealth indicators, water and sanitation access, and satisfaction with water service. The survey was administered electronically on password-protected mobile tablets by enumerators. Tablets were equipped with Open Data Kit (ODK) Collect, an open source program which allows offline data collection on a mobile device (25). A secure ODK compatible aggregation server was deployed for hosting the survey form and gathering the survey data. Submitted data were exported daily to ensure data quality (e.g., quality assurance using geocoded data to ensure households were within study area boundaries and spot checks to assess for missing survey data).

### Outcome variables

#### Water source

Respondents were asked to provide information on the main source of drinking water for members of the household. We classified households with a piped drinking water source located within the household or on the premises, with availability when needed, as “household connection”. Households without access to a piped water source at the household or on the premises were classified as “non-household connection”. These definitions were used to reflect criteria for safely managed drinking water according to service ladders of the Joint Monitoring Programme for Water Supply and Sanitation (JMP).(5)

#### Sanitation facilities

Respondents were asked to provide information on the main type of sanitation their household uses and whether the main type of sanitation is shared with people outside their household. Households with a basic sanitation facility – per JMP definitions – had an unshared facility. (5) Limited or unimproved sanitation facilities include the use of pit latrines without a slab or platform and could be shared between two or more households.

#### Satisfaction with water service

Respondents were also asked to report how often (never/sometimes/always) they are satisfied with overall service, water pressure, and water quality of their main source of drinking water. We asked respondents to report (yes/no) if there had been any time in the last month when the household did not have sufficient quantities of drinking water when needed. The responses to the water satisfaction questions related to services, pressure, and quality were recoded as binary, comparing those who were sometimes or always satisfied with their water provision (yes) to those who were never satisfied (no). A total satisfaction score was created by summing the individual binary scores (1=yes/satisfied, 0=no/unsatisfied), with the total score ranging between 0 and 4 for each household, and a higher score representing higher satisfaction.

### Predictor variables

#### Household demographics and assets and wealth indicators

We collected data on education of the primary caregiver, number of children under 5 years of age living in the household, and household density. Respondents answered ten standardized questions from the Simple Poverty Scorecard^®^ Poverty-Assessment Tool Mozambique, including questions on household size, materials, assets (Supplemental Material 3).(26) Each question’s answer choices correspond with a point total, and points are summed over all ten questions into a poverty score. We use this poverty score to compare consumption of assets across different households, both using it as a continuous score and categorizing it into quartiles.

#### Distance to water main

We calculated the distance of each respondent’s household to the water distribution main using a geocoded shapefile of the city’s water distribution system provided by FIPAG. A water main was defined by any pipe that had a diameter greater than 100 millimeters. The Euclidean distance between each survey respondent’s household and every water main pipe was calculated using the ‘sf’ package in R.(27) We then selected the minimum distance to a water main for each study participant for our analysis.

#### Statistical analysis

All statistical analyses were conducted using R statistical software (RStudio v. 1.3.1093). Bivariate analyses were conducted to describe the relationship between demographic variables and each of the outcome variables. We used unadjusted, logistic regression models to characterize associations between access to a household water connection or an unshared sanitation facility and sociodemographic variables (e.g., SES quartile) and water satisfaction responses.

We assessed whether there was statistically significant spatial heterogeneity in water satisfaction responses and household access to water and improved sanitation facilities. We applied a kernel density estimation approach to generate a spatial relative risk surface, which describes whether the density of a specific response in space is significantly different than the density of another response. Kernel density surfaces of bivariate density were generated for responses to each question using an over-smoothing, adaptive bandwidth approach. We then used the leave-one-out least-squares cross-validation (LSCV) risk function from the ‘sparr’ package (28) to select a jointly optimal, adaptive bandwidth for the kernel density surfaces from each question and used raster algebra to create the relative risk surface which contrasts the ratio of the numerator (e.g., at least sometimes satisfied) to the denominator (e.g., never satisfied). The resulting surfaces were mapped with *p*-value contours at an alpha level of 0.05, highlighting statistically significant spatial density of survey responses (R package: ‘spatstat’).(29) All maps were generated using the ‘tmap’ package.(30)

We used log-binomial regression to estimate the association between distance from water main pipe and having a household water connection. We used logistic regression to estimate the association between distance from water main pipe and satisfaction with water pressure, quality, service, and sufficiency. We assessed whether there was significant effect modification by onsite access to an improved water source on the relationship between distance to water main and satisfaction with water pressure. We used linear regression models to estimate the association between distance from the water main and total satisfaction score. Linear regression model assumptions were checked by analysis of partial plots, residual analyses, Q-Q plots, and variance inflation factors (R package: ‘car’); remedial measures were taken if applicable. Final models were stratified by significant interaction variables, when appropriate. Household density and SES score were included as confounders in each of the models based on a priori criteria.

## Ethics Statement

The study was approved by the Mozambique National Bio-Ethics Committee for Health (Ref: 105/CNBS/20) and the Institutional Review Board of Emory University (IRB#: CR001-IRB00098584, Atlanta, GA). In addition, we obtained permissions from local authorities, namely Beira municipality and municipal district administrations from study neighborhoods included in the study. Credential letters were issued to be presented in all sub-neighborhoods and household visited. Additionally, courtesy meetings between the study team and city health department were held. Recruitment and consent of subjects took place at the households. Prior to enrollment, study staff fully explained and carried out the consent process and documented the procedure. Subjects provided a signature to indicate consent. In the case of illiteracy of the subject, study staff verbally summarized the material with the subject, and the participants were required to mark the document with a thumbprint indicating their consent.

## Results

### Water and sanitation access by sociodemographic profile

A total of 773 (47.6%) households reported a household water connection (Table 1). Households in the wealthiest two SES quintiles (compared to the lowest quintile), those with a respondent with a high school or above education (compared to no formal schooling), and with more than eight people living in their household (compared to 1-4 people in their household) were more likely to have access to a household water connection. Those living with one or two children under five-years-old (compared to having no children) were less likely to have access to a household water connection.

**Table 1:** Socio-demographic profile of study recruits by household water connection in Beira, Mozambique.

|  | Household water connection* |  | OR (95% CI) <sup>†</sup> | p-value* |
| --- | --- | --- | --- | --- |
|  | Yes | No |  |  |
| **All respondents** | 773 (48%) | 836 (52%) |  |  |
| **Socioeconomic quartile** |  |  |  |  |
| Q1 (poorest) | 198 (26%) | 285 (34%) |  | Ref. |
| Q2 | 249 (32%) | 293 (35%) | 1.22 (0.95,1.57) | 0.11 |
| Q3 | 176 (23%) | 157 (19%) | 1.61 (1.22, 2.14) | <0.001 |
| Q4 (wealthiest) | 150 (19%) | 101 (12%) | 2.14 (1.57, 2.92) | <0.001 |

| Education level of primary caregiver |  |  |  |  |
| --- | --- | --- | --- | --- |
| No formal schooling | 68 (9%) | 78 (9%) |  | Ref. |
| Primary School | 114 (15%) | 131 (16%) | 1.00 (0.66,1.51) | 0.99 |
| Secondary school | 223 (29%) | 335 (40%) | 0.76 (0.53,1.10) | 0.15 |
| High school or above | 368 (48%) | 292 (35%) | 1.45 (1.01 ,2.07) | 0.04 |
| Children <5yrs living in household |  |  |  |  |
| 0 | 377 (49%) | 323 (39%) |  | Ref. |
| 1 | 244 (32%) | 328 (39%) | 0.64 (0.51,0.80) | <0.001 |
| 2 | 124 (16%) | 150 (18%) | 0.71 (0.54,0.94) | 0.02 |
| 3+ | 28 (4%) | 35 (4%) | 0.69 (0.41,1.15) | 0.15 |
| No. of people living in household |  |  |  |  |
| 1-4 | 264 (34%) | 346 (41%) |  | Ref. |
| 5-7 | 252 (33%) | 270 (32%) | 1.22(0.97, 1.55) | 0.09 |
| 8+ | 257 (33%) | 220 (26%) | 1.53 (1.20,1.95) | <0.001 |

*\*'Household water connection' was defined as those having piped water in own dwelling/yards.*
*†Odds ratio (OR) and 95% confidence intervals of an improved source of drinking water facilities among subjects' demographics were compared to those without using separate logistic regression models for each characteristic.*
*#P-values were obtained using chi-square tests*

A total of 862 (54%) households reported household access to a basic sanitation facility (Table 2). Households in the wealthiest socio-economic quintiles (compared to the lower quintile) and those with five or more people living in their household (compared to 1-4 people in their household) were more likely to have access to a basic sanitation facility. Those having one child under five-years-old (compared to having no children) were less likely to have access to a basic sanitation facility.

**Table 2:** Sociodemographic profile of study recruits by basic sanitation services in Beira, Mozambique

|  | Basic sanitation* |  | OR (95% CI) † | p-value# |
| --- | --- | --- | --- | --- |
|  | Yes | No |  |  |
| **All respondents** | 862 (54%) | 749 (46%) |  |  |
| Socioeconomic quartile |  |  |  |  |
| **Q1 (poorest)** | 240 (28%) | 248 (33%) |  | Ref. |
| **Q2** | 282 (33%) | 262 (35%) | 1.11 (0.87,1.42) | 0.39 |
| **Q3** | 175 (20%) | 155 (21%) | 1.17 (0.88,1.54) | 0.28 |
| **Q4 (wealthiest)** | 165 (19%) | 84 (11%) | 2.03 (1.48,2.79) | <0.001 |
| Education level of primary caregiver |  |  |  |  |
| **No formal schooling** | 76 (9%) | 71 (9%) |  | Ref. |
| **Primary School** | 123 (14%) | 119 (16%) | 0.97 (0.64,1.46) | 0.87 |
| **Secondary school (grades 5-8)** | 294 (34%) | 268 (36%) | 1.02 (0.71, 1.47) | 0.89 |
| **High school or above** | 369 (43%) | 291 (39%) | 1.19 (1.83, 1.70) | 0.35 |
| **Children <5yrs living in household** |  |  |  |  |
| **0** | 390 (45%) | 303 (40%) |  | Ref. |
| **1** | 291 (34%) | 286 (38%) | 0.79 (0.63, 0.99) | 0.04 |
| **2** | 141 (16%) | 136 (18%) | 0.81 (0.61, 1.07) | 0.13 |
| **3+** | 40 (5%) | 24 (3%) | 1.30 (0.76, 2.20) | 0.34 |
| **No. of people living in household** |  |  |  |  |
| **1-4** | 234 (27%) | 376 (50%) |  | Ref. |
| **5-7** | 295 (34%) | 229 (31%) | 2.07 (1.63, 2.63) | <0.001 |
| **8+** | 333 (39%) | 144 (19%) | 3.72 (2.88, 4.79) | <0.001 |

\*Basic sanitation facilities were defined as improved include flush/pour flush to piped sewer systems, septic tanks or pit latrines; ventilated improved pit latrines, composting toilets or pit latrines with slabs not shared with other households. Non-basic (i.e., limited or unimproved) sanitation facilities include pit latrines without a slab or platform, hanging latrines or bucket latrines shared between two or more households.
<sup>†</sup>Odds ratio (OR) and 95% confidence intervals of having improved sanitation facilities among subjects' demographics were compared to those without using separate logistic regression models for each characteristic.
\*P-values were obtained using chi-square tests

### Satisfaction with water services by household water access

For respondent-reported satisfaction with their water services, the total satisfaction mean score (0-4, with 4 being highest) was 3.45 (standard deviation (SD) 0.90) from respondents with a household water connection, and 3.44 (SD 0.83) for those without a household connection (Table 3). No differences were observed in satisfaction with water services, pressure, or quality between those with and without household water connections.

**Table 3:**
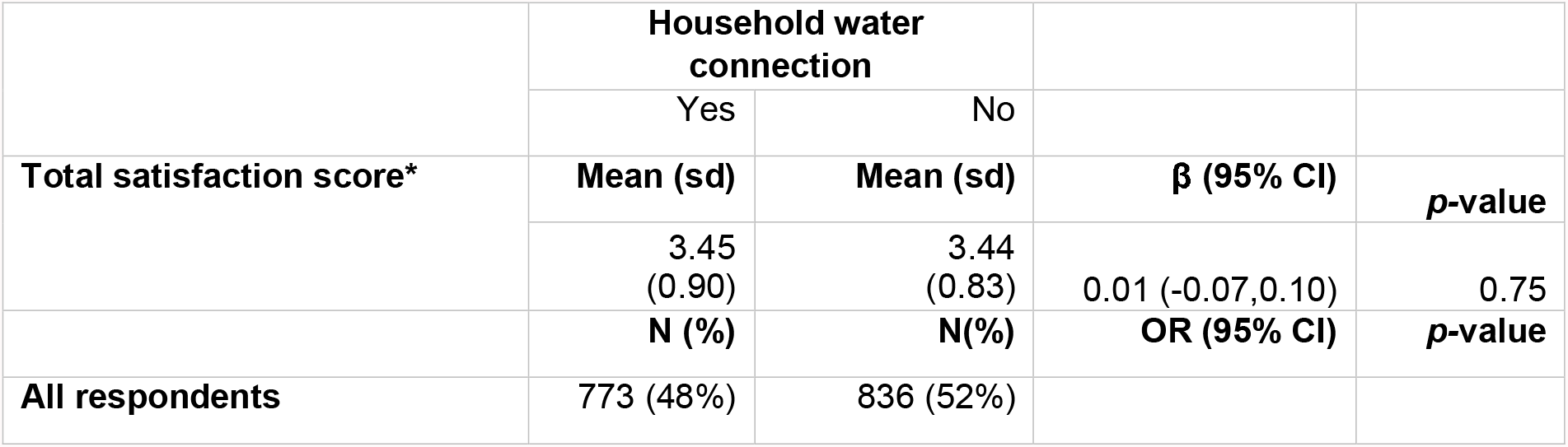
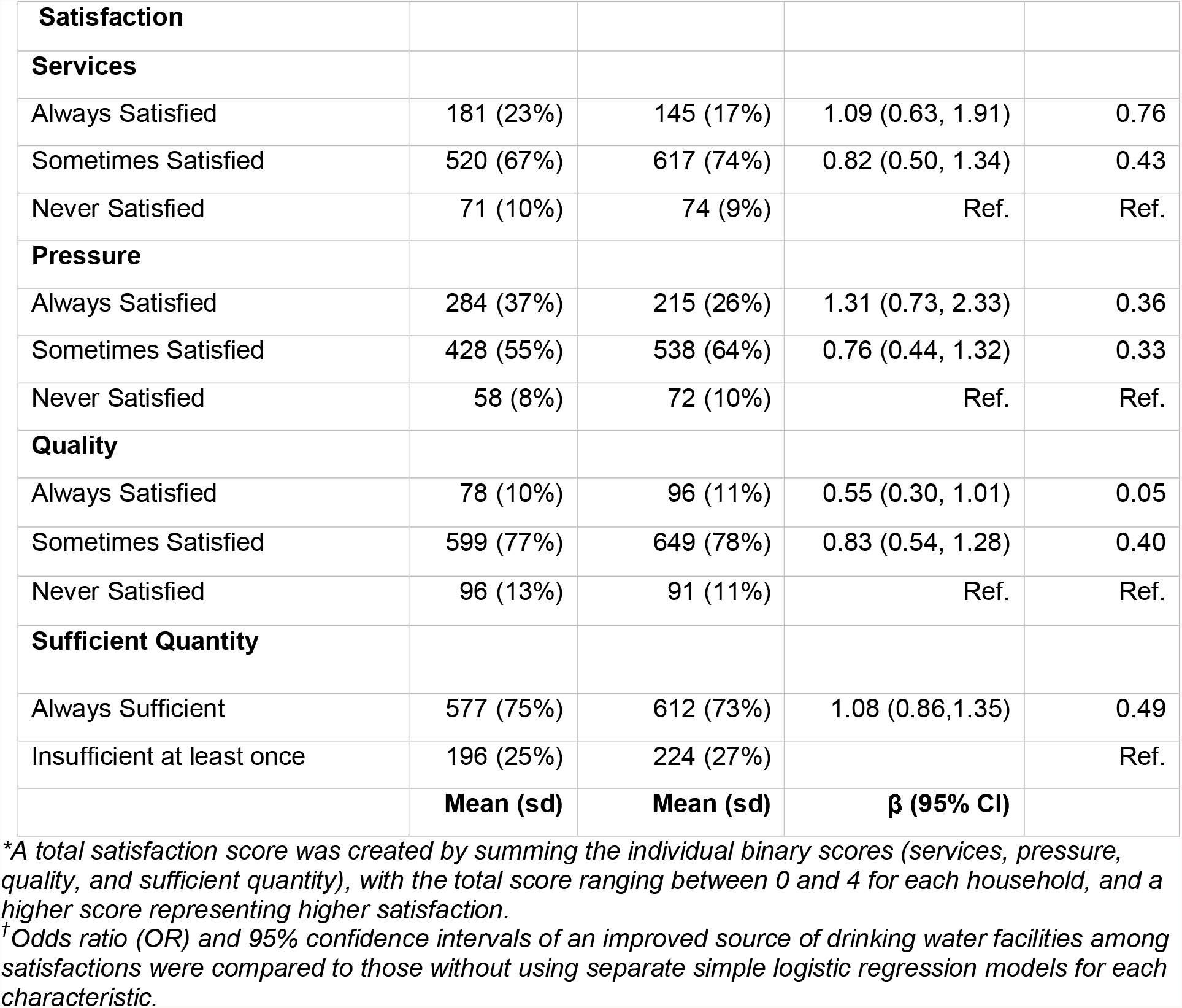
Frequency profile of respondent-reported satisfaction of water services in Beira, Mozambique

|  | Household water connection |  |  |  |
| --- | --- | --- | --- | --- |
|  | Yes | No |  |  |
| Total satisfaction score* | Mean (sd) | Mean (sd) | $\beta$ (95% CI) | p-value |
|  | 3.45 (0.90) | 3.44 (0.83) | 0.01 (-0.07, 0.10) | 0.75 |
|  | N (%) | N(%) | OR (95% CI) | p-value |
| All respondents | 773 (48%) | 836 (52%) |  |  |
| **Satisfaction** |  |  |  |  |
| **Services** |  |  |  |  |
| Always Satisfied | 181 (23%) | 145 (17%) | 1.09 (0.63, 1.91) | 0.76 |
| Sometimes Satisfied | 520 (67%) | 617 (74%) | 0.82 (0.50, 1.34) | 0.43 |
| Never Satisfied | 71 (10%) | 74 (9%) | Ref. | Ref. |
| **Pressure** |  |  |  |  |
| Always Satisfied | 284 (37%) | 215 (26%) | 1.31 (0.73, 2.33) | 0.36 |
| Sometimes Satisfied | 428 (55%) | 538 (64%) | 0.76 (0.44, 1.32) | 0.33 |
| Never Satisfied | 58 (8%) | 72 (10%) | Ref. | Ref. |
| **Quality** |  |  |  |  |
| Always Satisfied | 78 (10%) | 96 (11%) | 0.55 (0.30, 1.01) | 0.05 |
| Sometimes Satisfied | 599 (77%) | 649 (78%) | 0.83 (0.54, 1.28) | 0.40 |
| Never Satisfied | 96 (13%) | 91 (11%) | Ref. | Ref. |
| **Sufficient Quantity** |  |  |  |  |
| Always Sufficient | 577 (75%) | 612 (73%) | 1.08 (0.86, 1.35) | 0.49 |
| Insufficient at least once | 196 (25%) | 224 (27%) |  | Ref. |
|  | **Mean (sd)** | **Mean (sd)** | **β (95% CI)** |  |

\*A total satisfaction score was created by summing the individual binary scores (services, pressure, quality, and sufficient quantity), with the total score ranging between 0 and 4 for each household, and a higher score representing higher satisfaction.
<sup>†</sup>Odds ratio (OR) and 95% confidence intervals of an improved source of drinking water facilities among satisfactions were compared to those without using separate simple logistic regression models for each characteristic.

### Spatial heterogeneity of household water and sanitation access and consumer-reported water satisfaction

We identified statistical spatial heterogeneity in household access to water and basic sanitation (Figure 2A, 2B), as well as consumer-reported satisfaction with water services, pressure, quality, and sufficient quantity among our study participants within the city of Beira (Figure 2 C-F). The relative risk surfaces displayed in Figure 2 outline areas within the city that have significantly higher (green) or lower (red) density of access or lack of access, and satisfaction or lack thereof. Statistically high or low density of responses at an α-level of <0.05 are indicated by the contour line colored in white and blue, respectively. Some areas contain both statistically high and low access or satisfaction.

**Figure 2:**
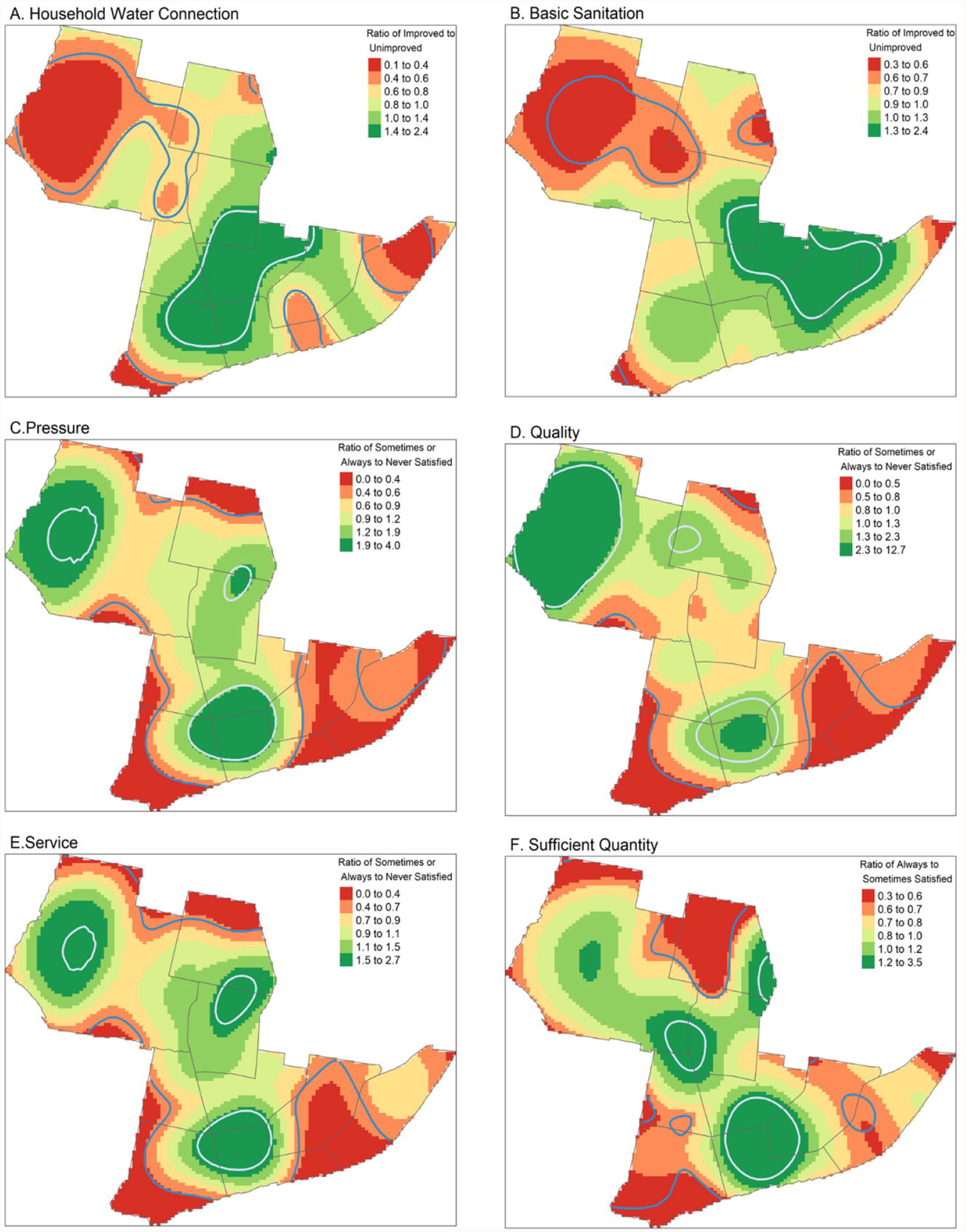
Relative risk surface of consumer-reported water satisfaction and improved water and sanitation access at the household P-value contours in blue and white indicate areas with statistically different high or low density of survey responses. A ratio value of 1 indicates when the probability of either response at a specific location are equal. A higher ratio indicates a higher probability of having household access to improved water or unshared sanitation services or being at least sometimes satisfied with the water services. An adaptive bandwidth selection was used to select the optimum bandwidth for each individual relative risk service.

Although the number of participants who had a household water connection and a basic sanitation facility was similar, those who had access to basic sanitation at the household were not always the same as those who had household water connections, indicated by the differences in the colored hotspots found in Figures 2A and 2B. Spatial patterns were similar across water satisfaction metrics (Figures 2C-F), but these metrics did not always overlap with household water connections (Figure 1A).

### Association of distance from water main with household access to water and satisfaction with water services

We found an inverse association between distance from water main and both access to a household water connection and satisfaction with water pressure, service, and sufficiency (i.e., as distance went up, satisfaction went down), controlling for household density and SES score. For every 100-meter increase in distance from a water main pipe, the prevalence of household access to an onsite water source was 13% lower (OR 0.87, 95% CI: 0.82, 0.92), controlling for only household density. The model did not converge with SES score included We also computed an odds ratio using logistic regression with SES score included, and found a similar effect estimate for the association between odds of a household having access to an onsite water source (OR: 0.82, 95% CI: 0.82, 0.92). Similarly, the odds of responding ‘sometimes’ or ‘always’ satisfied with water pressure – compared to ‘never’ - decreased by 20% for every 100-meter increase in distance from the closest water main pipe (OR: 0.80; 95% CI: 0.69, 0.94).

There was no effect modification by household access to water on the relationship between distance from water main and satisfaction with water pressure. The odds of responding ‘sometimes’ or ‘always’ satisfied with water service decreased by 18% for a 100-meter increase in distance from the water main (OR: 0.82, 95% CI: 0.70, 0.95). The odds of responding ‘always’ satisfied with water sufficiency-compared to the response of ‘insufficient at least once’-decreased by 21% for every 100-meter increase in distance from the water main (OR: 0.79, 95% CI: 0.71, 0.88). There was no association between distance from water main and satisfaction with water quality (OR: 1.02, 95% CI: 0.88, 1.19). SES and household density were significant confounders and subsequently included each of the models. We also observed an inverse association between total satisfaction score and distance from the water main, controlling for household density and SES score. For every 100-meter increase in distance from the water main, total satisfaction score was reduced by a factor of 0.08 (95% CI: -0.13, - 0.04) (Table 4).

**Table 4:** Assessment of the relationship between distance from water main pipe on household water access and consumer-reported satisfaction with water. Household density and SES score were included as covariates in each of the logistic regression models. SES was not included in the log binomial model for the association between distance from water main pipe and household water connection due to failed convergence. The coefficients correspond to a 100-meter increase in distance from the water main pipes.

| Outcome | Effect estimate | 95% CI | p-value |
| --- | --- | --- | --- |
| Household water connection | PR: 0.87<sup>#</sup> | 0.82, 0.92 | <0.01 |
| Water pressure satisfaction* | OR: 0.80<sup>†</sup> | 0.69, 0.94 | 0.01 |
| Water quality satisfaction* | OR: 1.02<sup>†</sup> | 0.88, 1.19 | 0.84 |
| Water service satisfaction* | OR: 0.82<sup>†</sup> | 0.70, 0.95 | 0.01 |
| Water sufficiency satisfaction* | OR: 0.79<sup>†</sup> | 0.71, 0.88 | <0.01 |
| Total satisfaction score | $\beta$: -0.08<sup>^</sup> | -0.13, -0.04 | <0.01 |

<sup>#</sup>Estimate is prevalence ratio computed using log binomial regression. This model included household density as a covariate, but not SES score.
<sup>†</sup>Estimates are odds ratios computed using logistic regression. Household density and SES score were included as covariates in each of these models.
\*Comparing responses of 'Always' or 'Sometimes' satisfied to the response of 'Never' satisfied
<sup>^</sup>Estimate is the coefficient computed using linear regression. Household density and SES score were included as covariates in this model.

## Discussion

We examined the heterogeneity in access to household water connections and sanitation, and satisfaction with water services in low-income peri-urban neighborhoods of Beira, Mozambique. By combining spatial statistical methods and regression analyses, we investigated whether locally-heterogenous factors such as distance to water mains influenced access to household water and basic sanitation facilities, and how access and water satisfaction varied. We found substantial spatial heterogeneity in access to and satisfaction with WASH services, even within a low-income, underserved area of the city. Distance to water mains was a key predictor of water access services and satisfaction, even over relatively short distances within neighborhoods.

We identified an inverse association between distance from water main pipes and access to a household water connection; the further the compound was from the water main, the less likely residents were to have onsite water access. This pattern was consistent as difference in distance increased (i.e., for a 500-meter increase in distance, odds of having onsite access to water decreased by 63%). While this result was expected given the principles of water distribution system engineering (i.e., household connections become more difficult to implement further from the distribution main), it is an important factor to consider in the infrastructure development process. Different approaches for water service delivery may be needed for those areas that are further from the water mains, particularly for those living in informal settlements (31, 32). The increase in urbanization in Mozambique and other LMICs has resulted in a disproportionate concentration in informal settlements, resulting in challenges related to water expansion and sanitation services.(18) Inequities in access to improved water system are closely linked to poverty, and continuing water insecurity further exacerbates already rising inequalities, resulting in prolonged public health concerns such as the spread of infectious diseases, malnutrition, limited economic development, and women and girls’ labor inequities.(9, 18, 33) Understanding predictors of access to an improved water source, such as distance to the water main pipes, can provide insight into water service expansion planning, which is a critical challenge in achieving sustainable development goal target 6.1, ensuring safe water access for all, particularly in urban areas. (17, 19, 34, 35)

We found a similar inverse relationship between distance from the water main and service satisfaction scores, in addition to significant spatial heterogenity in satisfaction with water services. One potential explanation for this result is that as the distance between the consumer from the water main increased, the length of the pipes that supply water to that consumer also increased. Longer service lines may have more connections, more stagnation, temperature fluctuations, and lower pressure, which could result in a lower satisfaction with water quality and service.(36, 37) While we did not see evidence for significant effect modification of water pressure, there might not have been enough variability in the binary water pressure responses to observe this effect. These results suggest that monitoring of water quality, pressure, and service on the most distal parts of the water system is important in improving the overall quality of the distribution system.

While water access is increasing in Mozambique, inequity remains a concern. We found statistical spatial heterogeneity in access to a household water connection and basic sanitation facilities within low-income neighborhoods of Beira. Previous estimates from the World Bank of water and sanitation coverage in Mozambique have only been applied on a regional scale, and assessed urban-rural disparities (5, 22). Such estimates usually describe rapid increases in WASH access in urban areas. Our results demonstrated that even city-wide estimates of water and sanitation coverage do not capture the local heterogeneity in access to these services. Such findings underpin discussions that global measures of coverage are largely overestimated, and highlight how little attention urban informal settlements receive relative to surrounding urban centres when it comes to development.(17, 20)

The unequal distribution of household connections to an improved water supply has important consequences. First, consumers who utilize a public standpipe tend to pay 3-4 times more for their water than those who have a private connection in their household or on their compound; these consumers face the additonal burden of time used and physical effort expended to collect their water from local sources.(16) An alternative to using a public standpipe is purchasing water from neighbors, which has been associated with lower satisfaction with water quality and a decrease in water availability.(16) Finally, water that is collected from outside of the household is subsequently stored and is subject to contamination with fecal material.(38-40) Thus, understanding how hetergeneous access to householdwater services influences behaviors around water usage is important to improve the control of waterborne diseases.

Households with water connections did not always have access to basic sanitation facilities. This phenomenon aligns with global reports of improved water access having increased at a greater rate than access to improved sanitation facilities.(41) Historically, funding agencies have been more willing to invest in water infrastructure than in sanitation.(42, 43) A potential reason for this finding is the differences in barriers to providing access to a household water connection and improved sanitation in informal settlements. Specifically, differences in access can be driven by the location of the settlement.(44) For example, a high water table, such as in Beira, can impede the installation of sanitation facilities such as pit latrines. (3, 23, 43) Further research should be directed towards the design of interventions to navigate the specific challenges related to the structure of informal settlements. Spatial maps, such as the ones we produced, could support planning and targeting of low-income and poorly served areas, recognizing the different engineering requirements of sanitation and water access.

In this study, we combined multiple analytical methods to explore neighborhood and sub-neighborhood heterogeneity of water access and satisfaction in Beira, Mozambique. Our study provided visual mapping of access to water and sanitation services to facilitate our understanding in the variability of urban coverage of access to improved water and sanitation facilities on a fine scale. Moreover, we were able to discern local heterogeneity of water access and satisfaction as a result of distance to water main. This study was also subject to limitations. We conducted the survey only in the low-income areas of Beira. Future research could measure and map city-wide access to water services and satisfaction to support more holistic and equitable planning. Survey questions related to water satisfaction were subjective and inherent to recall bias. Participants responses to water satisfaction questions may also be influenced by neighbors’ access to water services. Future studies could investigate the relationship between consumer satisfaction and perceived injustices or inequalities with water services. Additionally, standardized sampling methods could be applied to help avoid or limit needing recall information.

## Conclusion

Few studies have explored intra-neighborhood water and sanitation access in low-income urban neighborhoods. We found substantial spatial heterogeneity in access to and satisfaction with WASH services in low-income urban areas of Beira, even across small scales. Distance to water main was a key predictor of water access services and satisfaction. This finding highlights the challenges of providing equitable access to water in urban informal settlements, the need for infrastructural solutions that increase safe water access and pressure throughout neighborhoods, and the development of hybrid models of water service delivery that address heterogeneity in access even in areas that are theoretically served by piped water connections. Future research could explore solutions that allow for the manageable and sustainable expansion of service coverage without sacrificing quality. Understanding how heterogeneous access to improved water services influences water usage and behaviors can help control waterborne diseases and improve overall health in the growing urban areas of the world.

## Supporting information

STROBE Checklist for Cross-Sectional Studies

## Data Availability

Deidentified raw data and analysis code can be accessed on our project OSF (Open
Science Framework) site upon publication at this link: https://osf.io/2a963/. Geocoded
locations of study participants used to generate the maps will not be publicly available
due to ethical concerns and protection of our study participants.

## Abbreviations

LMICs: Low – and Middle-income Countries
WASH: water, sanitation and hygiene
SES: Socioeconomic status
FIPAG: Fundo de Investimento e Património do Abastecimento de Água
JMP: Joint Monitoring Programme for Water Supply & Sanitation
LSCV: Least-Squares cross-validation

## Acknowledgements

The authors would like to thank our study participants, who generously gave their time to participate in our survey. We thank WE Consult field supervisor (Mario Mungoi) and enumerators (Isabel Chiquel, Ricardina Timoteo, Maria Cazonda, Milauzia De Melo, and Marcelo Fernandes) who captured these data.

## Supplemental Figures and Tables

**Supplemental material 3:**
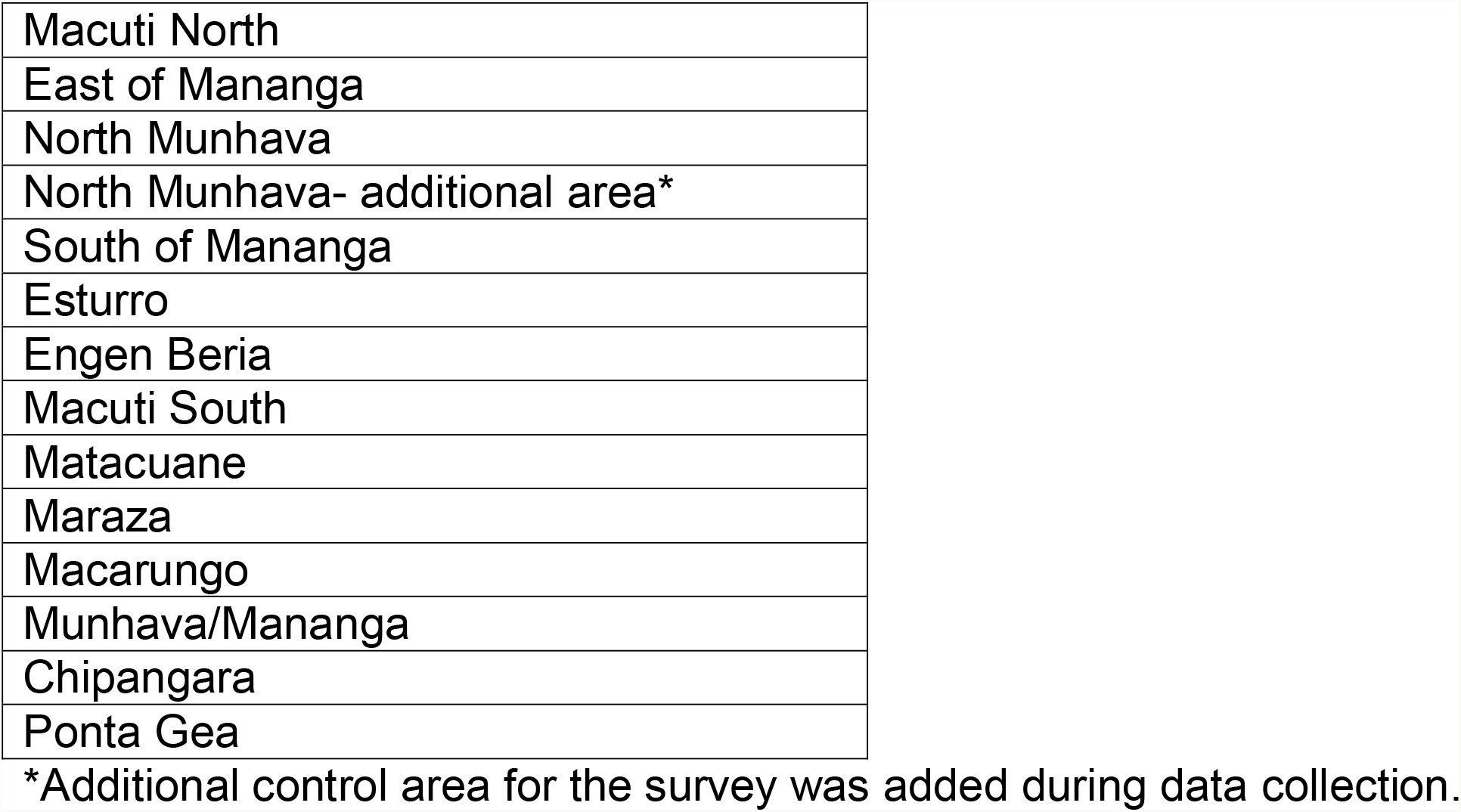
Study neighborhoods

**Supplemental material 4:** Standardized questions from the “Simple Poverty Scorecard® Poverty-Assessment Tool Mozambique,” which included questions on household size, materials, assets.

|  |
| --- |
| **HOUSEHOLD ASSETS & WEALTH INDICATORS** |
| How many bedrooms are in the house? |
| How many beds does this household have (single, double, bunkbeds, or for children)? |
| **Does your household/house have any of the following?** |
| A functioning bicycle |
| A functioning motorcycle |
| A functioning car/truck |
| Non-electric or electric clothes iron |
| Functioning freezer |
| A clock (mobile phone, wall, wrist, or pocket) |
| A radio, stereo system, or cassette player |
| A sofa |
| Grated windows |
| Does the house have a grated door? |
| What is the main source of energy for lighting in the residence? |
| Where is the kitchen located? |
| What is the main material of the floor of the residence (excluding kitchen and bathrooms)? |
| What is the main material of the walls of the residence? |

## References

1. Kallergis A. Water and Sanitation in the World’s Cities: Looking Ahead to 2050. 2013.

2. Programme UNHS. World Cities Report 2016: Urbanization and Development - Emerging Futures. Nairobi, Kenya: United Nations Human Settlements Programme (UN-Habitat); 2016. 262 p.

3. Sinharoy SS, Pittluck R, Clasen T. Review of drivers and barriers of water and sanitation policies for urban informal settlements in low-income and middle-income countries. Utilities Policy. 2019;60:100957.

4. Prüss-Ustün A, Wolf J, Bartram J, Clasen T, Cumming O, Freeman MC, et al. Burden of disease from inadequate water, sanitation and hygiene for selected adverse health outcomes: An updated analysis with a focus on low-and middle-income countries. Int J Hyg Environ Health. 2019;222(5):765–77.

5. (UNICEF) GWHOWatUNCsF. Progress on household drinking water, sanitation and hygiene 2000-2020: Five years into the SDGs. 2021. Contract No.: CC BY-NC-SA 3.0 IGO.

6. World Health O, United Nations Children’s F. Progress on drinking water, sanitation and hygiene: 2017 update and SDG baselines. Geneva: World Health Organization; 2017 2017.

7. Liu L, Johnson HL, Cousens S, Perin J, Scott S, Lawn JE, et al. Global, regional, and national causes of child mortality: an updated systematic analysis for 2010 with time trends since 2000. Lancet. 2012;379(9832):2151–61.

8. Troeger C, Blacker BF, Khalil IA, Rao PC, Cao S, Zimsen SRM, et al. Estimates of the global, regional, and national morbidity, mortality, and aetiologies of diarrhoea in 195 countries: a systematic analysis for the Global Burden of Disease Study 2016. The Lancet Infectious Diseases. 2018;18(11):1211–28.

9. UNESCO E. The United Nations world water development report 3: Water in a changing world. UNESCO Earthscan; 2009.

10. Collaborators GMaCoD. Global, regional, and national life expectancy, all-cause mortality, and cause-specific mortality for 249 causes of death, 1980-2015: a systematic analysis for the Global Burden of Disease Study 2015. Lancet. 2016;388(10053):1459–544.

11. Pru□ss-U□stu□n A, World Health O. Safer water, better health : costs, benefits and sustainability of interventions to protect and promote health. / Annette Pru□ss-U□stu□n … [et al]. Geneva: World Health Organization; 2008.

12. Water S, Hygiene and Health, WHO Headquarters (HQ), WHO Worldwide. Water, sanitation, hygiene, and waste management for SARS-CoV-2, the virus that causes COVID-19. 2020:15.

13. Fund UNCs. Global Framework for Urban Water, Sanitation and Hygiene. New York: UNICEF; 2019.

14. Clark H, Coll-Seck AM, Banerjee A, Peterson S, Dalglish SL, Ameratunga S, et al. A future for the world’s children? A WHO–UNICEF–Lancet Commission. The Lancet. 2020;395(10224):605–58.

15. Santoro JSaJ. Urbanization in Sub-Saharan Africa: Meeting Challenges by Bridging Stakeholders: Center for Strategic and International Studies (CSIS); 2018.

16. Luengo M, Banerjee S, Keener S. Provision Of Water To The Poor In Africa : Experience With Water Standposts And The Informal Water Sector: The World Bank; 2010. 65 p.

17. Nganyanyuka K, Martinez J, Wesselink A, Lungo JH, Georgiadou Y. Accessing water services in Dar es Salaam: Are we counting what counts? Habitat International. 2014;44:358–66.

18. Hulya D, Simon AR. Access to water in the slums of the developing world. 2009. Contract No.: 57.

19. Dos Santos S, Adams EA, Neville G, Wada Y, de Sherbinin A, Mullin Bernhardt E, et al. Urban growth and water access in sub-Saharan Africa: Progress, challenges, and emerging research directions. Sci Total Environ. 2017;607-608:497–508.

20. Satterthwaite D. Missing the Millennium Development Goal targets for water and sanitation in urban areas. Environment and Urbanization. 2016;28(1):99–118.

21. Tincani L, Ross, I., Zaman, R., Burr, P., Mujica, A., Evans, B. Regional assessment of the operational sustainability of water and sanitation services in Sub-Saharan Africa. Oxford, UK: Oxford Policy Management; 2015.

22. Bank W. Water Supply and Sanitation in Mozambique: Turning Finance into Services for 2015 and Beyond. The World Bank; 2012.

23. Group WB. Findings of the Mozambique Water Supply, Sanitation, and Hygiene Poverty Diagnostic. Washington, DC: World Bank; 2018.

24. UNICEF, editor Sanitation in Small Towns : Experience from Mozambique 2016.

25. Hartung C, Lerer, A., Anokwa, Y., Tseng, C., Brunette, W., & Borriello, G. Open data kit: tools to build information services for developing regions. Proceedings of the 4th ACM/IEEE international conference on information and communication technologies and development. 2010:1–12.

26. Schreiner M. Simple Poverty Scorecard Poverty-Assessment Tool Mozambique. Swiss Development Corporation/Microfinance Risk Management, LLC. 2013:127.

27. Pebesma E. Simple Features for R: Standardized Support for Spatial Vector Data. R Journal. 2018;10:439–46.

28. Davies TM, Marshall JC, Hazelton ML. Tutorial on kernel estimation of continuous spatial and spatiotemporal relative risk. Stat Med. 2018;37(7):1191–221.

29. Turner R, Baddeley A. SPATSTAT: an R package for analyzing spatial point patterns. Journal of Statistical Software. 2005;12.

30. Tennekes M. tmap: Thematic Maps in R. 2018. 2018;84(6):39.

31. Shah M, Fabrizio, N., Clark, B., & Veitch, J. Improving Service Delivery to Informal Settlements through Data Management. Cape Town: Worcester Polytechnic Institute; 2016.

32. Bank W. Unsettled: water and sanitation in urban settlement communities of the Pacific. Situation Report. 2015 19 Nov 2015.

33. Angoua ELE, Dongo K, Templeton MR, Zinsstag J, Bonfoh B. Barriers to access improved water and sanitation in poor peri-urban settlements of Abidjan, Côte d’Ivoire. PLoS One. 2018;13(8):e0202928–e.

34. Mora-Rodríguez J, Delgado-Galván X, Ortiz-Medel J, Ramos HM, Fuertes-Miquel VS, López-Jiménez PA. Pathogen intrusion flows in water distribution systems: according to orifice equations. Journal of Water Supply: Research and Technology-Aqua. 2015;64(8):857–69.

35. Transforming our world: the 2030 Agenda for Sustainable Development. 21 October 2015.

36. Council NR. Introduction. Drinking Water Distribution Systems: Assessing and Reducing Risks. Washington, DC: The National Academies Press; 2006.

37. Reynolds KA. Vulnerabilities of the Drinking Water Distribution System: Water Conditioning & Purification International Magazine; 2007 [

38. Zuin V, Ortolano L, Alvarinho M, Russel K, Thebo A, Muximpua O, et al. Water supply services for Africa’s urban poor: the role of resale. Journal of Water and Health. 2011;9(4):773–84.

39. Holcomb DA, Knee J, Sumner T, Adriano Z, de Bruijn E, Nalá R, et al. Human fecal contamination of water, soil, and surfaces in households sharing poor-quality sanitation facilities in Maputo, Mozambique. Int J Hyg Environ Health. 2020;226:113496.

40. Chalchisa D, Megersa M, Beyene A. Assessment of the quality of drinking water in storage tanks and its implication on the safety of urban water supply in developing countries. Environmental Systems Research. 2017;6(1):12.

41. Programme UND. The vast deficit in sanitation. Human Development Report 2006: United Nations; 2006.

42. Isunju JB, Schwartz K, Schouten MA, Johnson WP, van Dijk MP. Socio-economic aspects of improved sanitation in slums: A review. Public Health. 2011;125(6):368–76.

43. Cairncross S, Bartram J, Cumming O, Brocklehurst C. Hygiene, Sanitation, and Water: What Needs to Be Done? PLOS Medicine. 2010;7(11):e1000365.

44. Chaplin SE. Indian cities, sanitation and the state: the politics of the failure to provide. Environment and Urbanization. 2011;23(1):57–70.

