## Supplementary figures and images for "SPATIAL HETEROGENEITY OF NEIGHBORHOOD-LEVEL WATER AND SANITATION ACCESS IN INFORMAL URBAN SETTLEMENTS: A CROSS-SECTIONAL CASE STUDY IN BEIRA, MOZAMBIQUE"

### Supplemental Figure 1

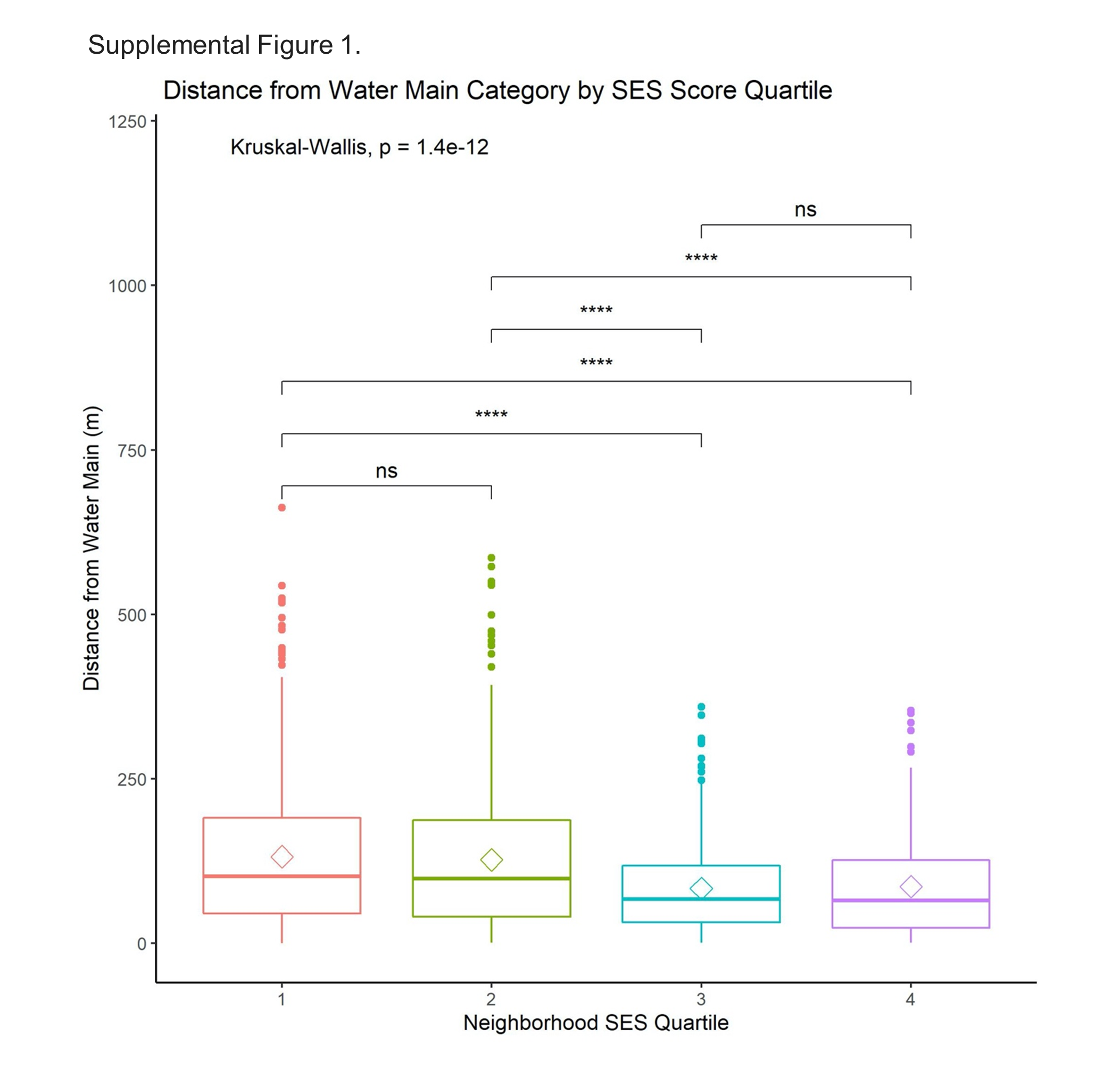

### Supplemental Figure 2

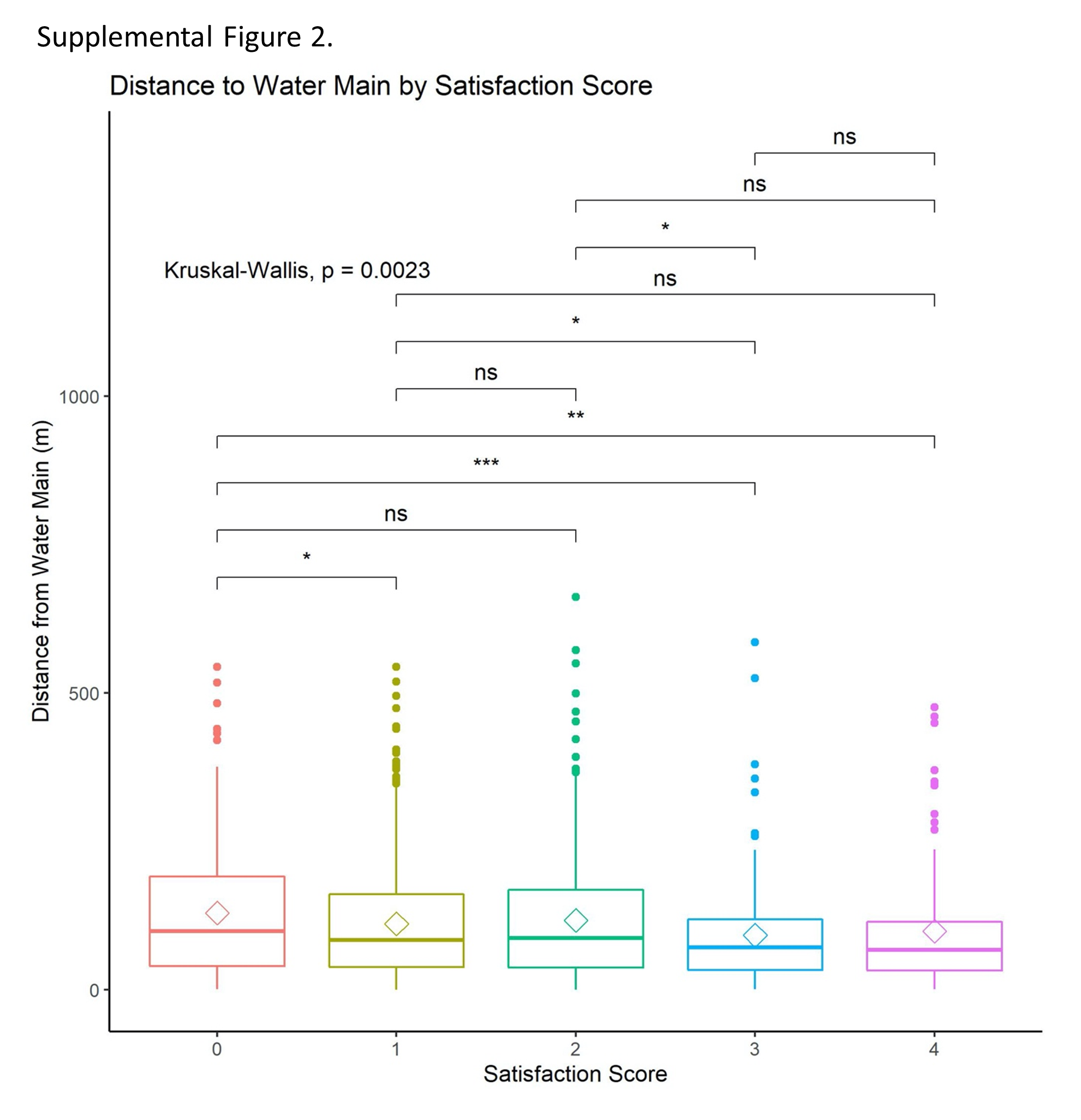
